# Methods for a population-based Comprehensive Eye care Workload Assessment (CEWA) Study in Southern India

**DOI:** 10.1101/2023.01.01.23284101

**Authors:** Ashok Vardhan, Vinoth Kumar Rajendran, Sanil Joseph, Lakshmanan Pooludaiyar, Dipankar Datta, Astrid E Fletcher, Thulasiraj Ravilla

## Abstract

Eye care programs, in developing countries, are often planned using the cataract surgical rate (CSR) targets - retrospectively estimated from Rapid Assessment (RAAB) surveys. A limitation of this approach is that it ignores the annual overall eye care requirements for a given population. Moreover, targets set are arbitrary, often influenced by capacity rather than need. To address this lacunae, we implemented a novel study design to estimate the annual need for comprehensive eye care in a 1.2 million population.

We conducted a population-based longitudinal study in Theni district, Tamil Nadu, India. All permanent residents of all ages were included. We conducted the study in three phases, (i) household-level enumeration and enrolment, (ii) basic eye examination (BEE) at household one-year post-enrolment, and (iii) assessment of eye care utilization and full eye examination (FEE) at central locations. All people aged 40 and above were invited to the FEE. Those aged less than 40 years were invited to the when FEE if indicated.

We conducted three pilot studies to test the study and clinical examination protocols. In the main study we enrolled 24,327 subjects (58% aged below 40 years and 42% aged 40 years and above). Of those less than 40 years, 72% completed the BEE, of whom 20% were referred for FEE and 70% of people aged ≥40 years underwent a comprehensive eye examination at central location.

Our study design provides appropriate long-term public health intervention planning, resource allocation, efficient delivery of care, and designing of eye care services for resource-limited settings.

## Introduction

Improvements in socioeconomic status and the accessibility of eye care have resulted in decrease in the prevalence of visual impairment (VI) and blindness in recent years.^[1]^ Joint efforts of various stakeholders under the Vision 2020 The Right to Sight initiative have also contributed to this.^[2]^ However, age-related eye diseases will continue to affect an increasing number of people globally, as the number of individuals 60 years and older is expected to double over the next 30 years.^[1,3,4]^ Unpublished routine hospital data from Aravind Eye Hospital (AEH), Theni shows that 50% of outpatient services are accessed by people aged below 40 years and 54% of those who underwent cataract surgery during one year, had a pre-operative visual acuity (VA) ≥ 6/18 in the better eye and 12% had pre-operative VA ≥ 6/18 in the operated eye. Such earlier intervention, is likely due to increased functional vision needs, predominantly owing to an increase in literacy and use of technology such as mobile phones and television. This changing trend in VA thresholds for cataract surgery,^[5]^ seems to influence patient-driven demand for eye care services in India and other low- and middle-income countries. These observations, along with a very large proportion of the population with uncorrected presbyopia,^[6]^ underline the need to scale up efforts to address the burden of vision-related ailments at all levels.^[7]^

There exist a number of shortcomings in the current planning of eye care services. First, in many developing countries planning is driven by arbitrary CSR targets - based on backlog estimates derived from Rapid Assessment (RAAB) surveys ^[8]^ that by design, focus only on estimating prevalence. Second, most eye care programmes are geared towards patients who seek care; primarily at a stage when marked deterioration in vision has set in. As such, the number of people who do not seek care (unmet need) for their vision-limiting eye diseases greatly exceeds those who do (met need).^[9]^ Third, older individuals are likely to be affected by more than one eye problem and require comprehensive screening and management that can address multiple eye diseases.^[10]^ This warrants a change in our approach, in order to deliver holistic care, instead of designing programmes that focus on specific eye conditions or limited to those seeking care. We need to set targets based on need in the community.

These factors pose a major challenge for both eye care providers and policymakers and demand the establishment of an efficient service delivery model to reduce the current backlog of VI, focus on preventing chronic conditions and successful management of the increasing eye care demands emerging from population trends.

Given this scenario, there is a need for a study design that can estimate the eye care workload in a given community so as to enable eye care providers to plan services recognizing resource requirements. This requires reliable data on the current extent and pattern of eye care utilization, prevalence and causes of blindness and VI in all age groups. In order to address this gap, we developed a comprehensive eye care workload assessment (CEWA) methodology; a novel survey approach that can potentially estimate the need for eye care service in a given population.

The primary objective of this study was to estimate a twelve-month workload for providing comprehensive eye care for the priority eye conditions under VISION 2020 – The Right to Sight, India, namely: cataract, refractive errors, diabetic retinopathy, glaucoma, childhood blindness and low vision.^[2]^ The secondary objectives were (1) to identify the barriers and enablers for use of eye care services and (2) to report the prevalence of blindness and VI due to various eye diseases.

## Methods

We conducted a population-based longitudinal study between August, 2015 and April, 2018 in Theni district, Tamil Nadu, India (Figure-1). The study was conducted in three phases, (i) household-level enumeration and enrolment, (ii) preliminary eye examination at households (HH) one-year post-enrolment and (iii) detailed eye examination at central cluster locations. (Figure-2).

**Figure 1:**
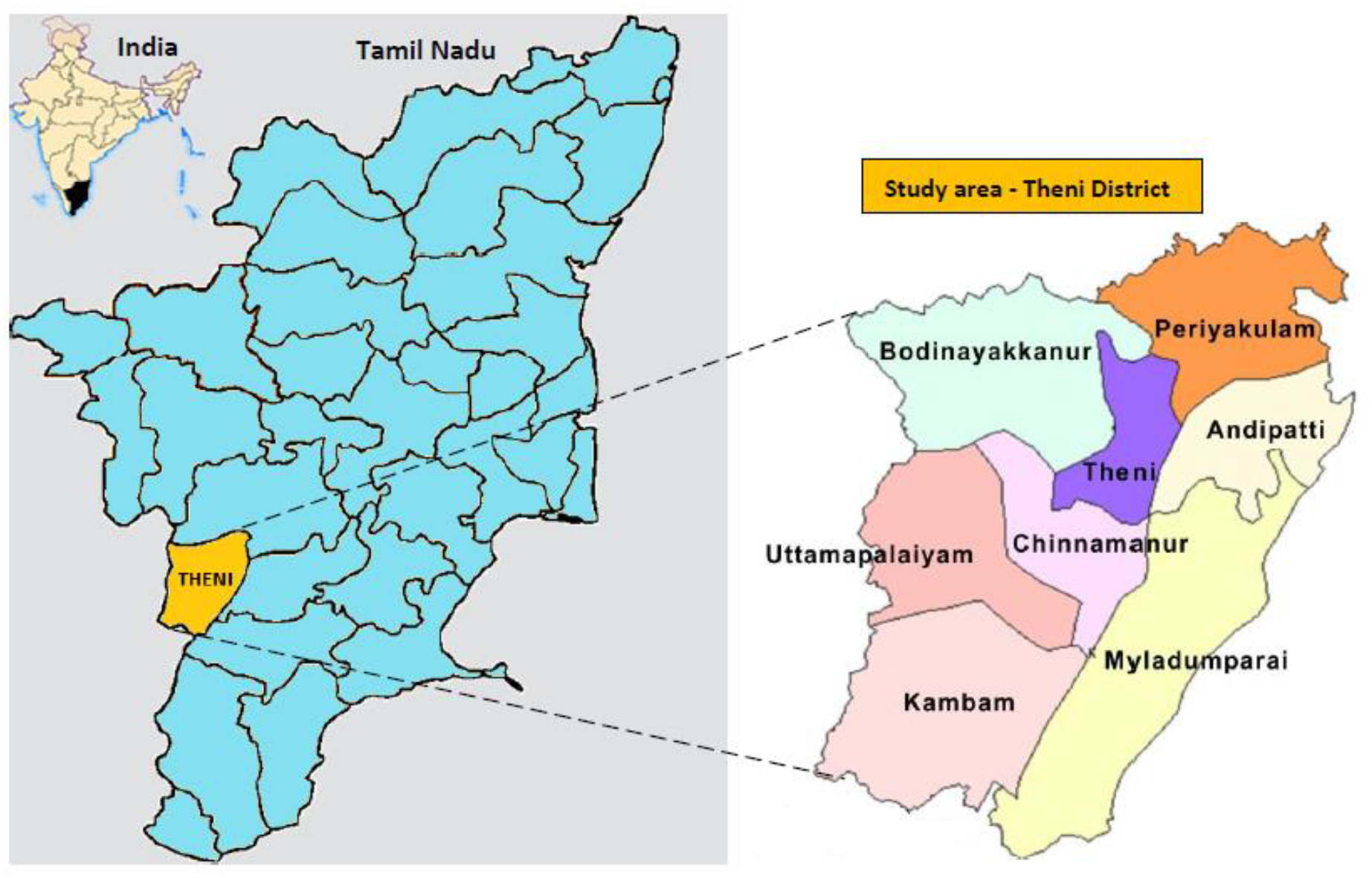
Study area map

**Figure 2:**
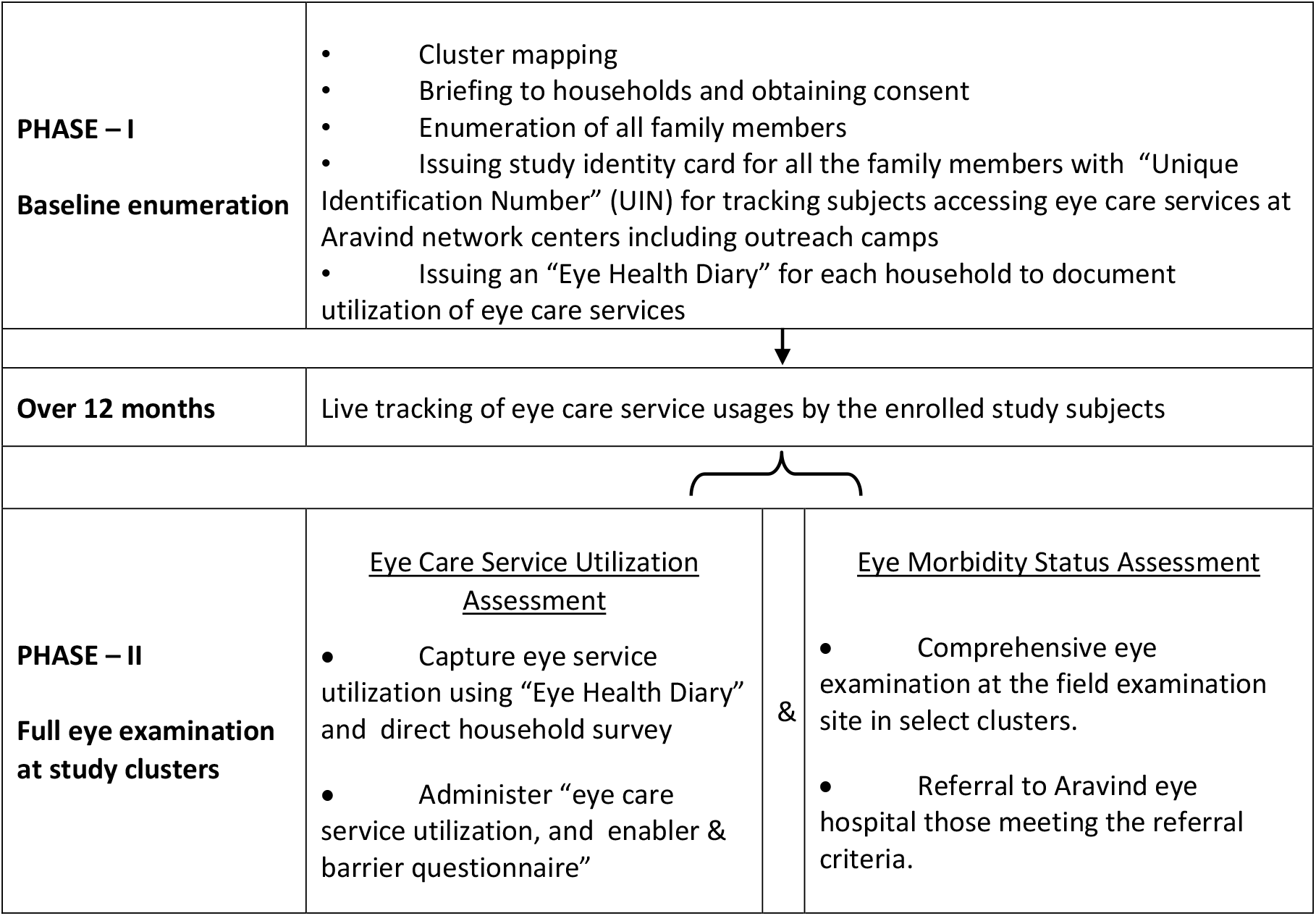
Study design

The study protocol was approved by the Institutional Review Board (Reg. No. ECR/182/Inst/TN/2013 dated 20.04.2013) of Aravind Eye Hospital, Madurai. We obtained written-informed-consent from all subjects; parents/guardians gave consent on behalf of subjects <18 years.

### Study area

Theni district has a population of 1.2 million ^[11]^ with a comparable mix of urban (55%) and rural (45%). According to the *Tamil Nadu State Blindness Control Society*, Theni has a district CSR of 9,781 (2015-16). AEH has a well-developed eye care network in the district consisting of 7 primary eye care centres, 1 urban eye clinic and a base hospital providing secondary and limited tertiary level services. Additionally, regular comprehensive screening eye camps are conducted throughout the year across the district. ^[12]^ AEH Theni, performs over 9,300 cataract surgeries annually on patients from Theni district. Using AEH’s contribution to CSR as a surrogate indicator, it is reasonable to assume that 80% of the eye care services in the district are provided by the AEH network. Since most patients receive eye-care from AEH, it offers the potential to prospectively track eye care usage by this population.

### Sample size and sampling procedure

The sample size was calculated assuming a 2.0% prevalence of blindness among people aged ≥40 years ^[9]^ and an error bound (precision) of 20% at 95% confidence interval. This was adjusted for the homogeneity within HH and cluster levels using a design effect of 1.5. Allowing for an approximate 15% non-response due to refusals, physical illness, death or migration (i.e. 85% response rate), the sample required was 8500 subjects aged ≥40 years. To enrol 8,500 subjects with >40 years, 25,000 persons of all age groups were required, since 40+ accounts for 35% of total population.^[11]^ A random cluster sampling method with probability proportional to size (PPS) was used. Sampling was done at the HH level from selected clusters, covering all age groups. With each cluster population ranging from 800 to 1,000, we randomly selected 34 clusters (17rural+17urban) from a sample frame of 1,454 clusters (urban-787, rural-667).

### Inclusion and exclusion criteria

All permanent residents of all ages, living in the same HH at least for the past 6 months, as told by the informant during enrolment, were included. Terminally or severely ill patients and mentally disabled persons were excluded from the study.

### Recruitment

Prior to baseline enumeration, cluster mapping was done with the help of volunteers and local leaders. After obtaining informed-consent, all members of the HH in the randomly selected clusters were enrolled into the study and were assigned a unique ID. An individual study identity card with a unique identification number was given to each member of the HH. A study ID number was also assigned to each HH and a sticker printed with HH ID was stuck on the main door. Photographs of the front view of the enrolled houses were taken for future reference. Data entry during enumeration was done real-time in the field and a printed ‘eye health diary’ with details of family members was issued to each HH. The eye health diary was designed to record details on eye care utilization of the subjects during the following twelve months. The participants were requested to present their eye health dairy whenever they accessed eye care services and request the the care provider to fill in a few basic details such as date, chief complaints, diagnosis and treatment advice.

### Tracking of eye care utilization

In addition to the ‘eye health diary’, we adopted two processes for tracking the utilization of eye care services following the enrolment: (1) live-tracking (post-baseline-enumeration) and (2) retrospective-tracking (post-morbidity-assessment survey). *Live-tracking* began from the date of enrolment for each cluster and lasted until the morbidity assessment survey. Tracking happened throughout the district in all nine AEH centres and in outreach camps conducted by AEH. Appropriate training was provided to all vision technicians in the primary eye care centres and the urban clinic. All study participants’ unique IDs along with their respective mobile phone numbers were uploaded in the hospital management system so as to get an alert whenever a study participant registered at any AEH centre. A poster displaying the study title and study village names was put up on the registration counter of all the AEH centres to remind the subjects to report to the registration staff regarding their participation in the study. No financial or other incentives were provided to the participants so as not to skew the utilization pattern. One staff member was posted at the registration counter at the base hospital specifically for tracking study participants reporting to the hospital and another staff member with enumeration-database installed in a laptop was posted at all outreach camps in the proximity of study clusters. Retrospective tracking was done based on self-reporting by the participants in response to the eye healthcare utilization questionnaire at the time of the final morbidity-assessment survey.

### Morbidity Assessment

We adopted two different approaches to assess morbidity at the community level (a) Basic Eye Examination for those aged below 40-years at HHs and (b) Full Eye Examination for those ≥40 years, at central cluster locations and those referred from HH after the Basic Eye Examination.

### Pilot study

We conducted three pilot studies to test out the examination protocols and to assess the response rates at the final morbidity-assessment survey. Since the response rate was less than 65%, in the first two pilots, a third pilot was conducted. To improve the response rates free medicines and spectacles were offered to those requiring them and we also extended the screening hours so as to make it more convenient to the participants.

### Basic Eye Examination (BEE)

BEE was conducted at HHs for each cluster which took about 2-3 days depending upon cluster size. For those below 40-years, the VA and external eye examination (Table-1) was performed by two trained ophthalmic technicians and two ophthalmic assistants, respectively. Subjects meeting the referral criteria (Figure-3) were referred for full eye examination (FEE). All other subjects, ≥40 years were directly referred for FEE. A minimum of three visits were made to examine ‘non-contactable’ subjects; if unsuccessful, then a referral card was hand-delivered to their HH members/neighbours, requesting them to directly come to FEE centre in their locality.

**Table 1:** Screening protocol for the basic eye examination (BEE) performed at household level.

| Eye examination protocol for subjects less than 40 years old |
| --- |
| • Children <5 years were examined with standard VA tests corresponding to their age (Figure-3).<br>• For subjects between 5-39 years, VA was examined using PEEK visual acuity app (Version 3.3, Peek Vision Ltd, London, UK) installed in a smartphone.<br>• Torch light examination was done to identify common eye pathologies like Bitot's spot, congenital cataract, ptosis, buphthalmos, squint, nystagmus, white reflex, red eye, etc.<br>• Besides, the subjects were asked a few (Yes/No) questions regarding the use of spectacles, any disturbing eye problem and access to eye care in the previous 12 months.<br>• 'Disturbing eye problems' were defined as 'defective vision, injury, chronic watering or discharge lasting for 2 days, and redness or pain or swelling in either eye for three or more days. When the answer was 'Yes' to any of these questions, or when the presenting VA (PVA) was worse than 6/9.5 in either eye, the subject was referred for a full eye examination to be held within study cluster locations. |

### Full eye examination

Following BEE, FEE was conducted at a community hall within the cluster for all subjects ≥40 years and those <40 years who were referred from HH screening. The FEE clinical team consisted of 17 staff. After obtaining informed consent, a staff member administered an “eye healthcare utilization questionnaire” to identify the participant’s usage of eye care services in the preceding twelve months. The questionnaire included questions relating to ocular conditions, eye care provider approached, treatment advice and information about uptake of the advised treatment. For subjects, who had not accessed any eye service in previous twelve months, the time of their last eye exam and the service provider were obtained. The study ophthalmologist cross verified the interview responses while performing the eye examination and finalising the probable diagnosis. Subsequently, the participant underwent clinical examinations that included VA assessment, subjective refraction, intra-ocular-pressure measurement, systemic disease evaluations, capillary blood sugar test, HbA1c, preliminary eye examination, fundus photography and dilated slit lamp examination (Table-2). The process flow of clinical examination is shown in Figure-3.

**Table 2:** Screening protocol for full eye examinations (FEE) held at central cluster locations

| Eye examination | Measurement protocol |
| --- | --- |
| Visual acuity (VA) test | The VA was tested in each eye separately with the subject wearing habitual spectacles (if any) using the Early Treatment of Diabetic Retinopathy Study (ETDRS) tumbling E chart at a distance of 4 m and recorded as Snellen equivalent ($\geq 4$ of 5 letters correctly identified in each row). The ETDRS charts were made of non-reflective white polystyrene material and were installed in a retro illuminated box (2 X 2 feet). The vision testing started with the top of the chart and continued until a line was reached where more than half the letters (for example, two of four, three of five) were read incorrectly or the patient read all letters on the chart. Following distance VA, unaided near vision was tested using ETDRS near vision chart held at 40 cm distance. All the tests were performed for the right eye first followed by the left eye. |
| Subjective refraction | All subjects underwent objective refraction using streak retinoscope (BETA 200, HEINE, GERMANY) followed by subjective refraction. The best-corrected VA (BCVA) was captured after providing the best possible subjective correction to the subject's eyes. Vision with pinhole was measured for subjects with BCVA<6/9 and also for subjects who did not have a refraction because of red eye. |
| Intra-ocular pressure (IOP) measurement | IOP was measured with Perkin's applanation tonometry (MK3, Clarke Ophthalmic, Essex, UK) by a trained ophthalmic assistant. Subjects were explained about the procedure and 0.5% proparacaine was used to anesthetize the eye and 1% fluorescein strip wetted with 2% hypromellose solution was used to stain the eyes. |
| Systemic disease evaluation | Subjects aged 40 years and above underwent systemic disease evaluation. A trained nurse obtained consent and then interviewed the subjects to capture any known systemic conditions like diabetes, hypertension, cardiac diseases and bronchial asthma. If present, the treatment details for the relevant conditions were captured. |
| Blood Pressure (BP) measurement | All BP measurements were made using HEM-7121OMRAN (OMRAN Healthcare Singapore) automated BP instrument. A single BP recording was taken with subject seated in a chair and with the BP instrument placed approximately at a table that was at the level of subject's heart. |
| Capillary Blood Sugar Test | Capillary blood sugars were measured using Accutome glucometer (Model NC, ACCUTOME) and recorded as "Random" (>120 min after any food/drink consumption) or "postprandial blood sugar (PPBS)" (90 min to 120 min after any food/drink consumption) or "Fasting Blood Sugar" (fasting for at least 8 hrs). Subjects, who had consumed anything within the past 90 minutes, were temporarily postponed from undergoing blood sugar tests until the time had crossed at least 90 minutes to check PPBS. |
| HbA1c Test | In a previously non-diabetic, if the RBS (Random Blood Sugar) $\geq 140\text{mg\%}$ or PPBS $\geq 180\text{mg\%}$, then those subjects were made to undergo HbA1c measurements. HbA1c measurements were offered at the same site using Alere Afinion AS100, (Alere Inc. Waltham, Massachusetts, United States) capillary HbA1c measuring device. HbA1c values $>6.5$ was considered diabetic. |
| Preliminary eye examination | Following refraction and systemic diseases evaluation, the subjects underwent a preliminary eye examination by an ophthalmologist. Four ophthalmologists were posted for this full eye screening. The ophthalmologist did a torch light examination followed by examination using a slit lamp (SL-2G, Topcon Corporation, Japan). Anterior chamber depth was measured using Van Herrick's grading. Presence of pterygium, corneal opacity, PXF at pupil etc. and other anterior segment pathologies were documented. The undilated fundus was visualized using 90 dioptre lens. 2 mirror gonioscopy (O2M, two mirror gonio diagnostic lens, OCULAR INSTRUMENTS, Inc, BELLEVUE, WA, USA) was done for patients who were already known to have glaucoma or glaucoma suspects. The criteria followed were a) IOP$>21\text{mm Hg}$ or b) Van Herrick grade $\leq 2$ c) Vertical CDR $>0.6$ or CDR asymmetry $>0.2$ d) Abnormal anterior segment deposits such as pseudo exfoliation, pigment dispersion syndrome e) peripheral anterior synechiae or other findings consistent with secondary glaucoma and the patients were dilated with 1% tropicamide eye drops. |
| Pupil dilation | All subjects $\geq 40$ years were counselled for dilated eye examination. Subjects who had occludable angle, allergy to tropicamide eye drops and subject not willing for dilatation were not dilated. |
| Fundus photography | After dilatation, the patients underwent fundus photography using fundus camera (V 3.2.9.4256, BOSCH, Stuttgart, Germany). Minimum 2 fundus photos were taken for each eye. Disc-centred and macula-centred pictures were taken. |
| Slit lamp examination after dilation | The dilated fundus was visualized using 90 dioptre lens. The cataract was graded using World Health Organization (WHO) cataract grading system (13). In eyes with pseudophakia, the PCO status, Yag capsulotomy status was noted. PXF over lens was also noted. The vertical cup disc ratio, the status of the macula, and background retina were recorded. |

**Table 3:**
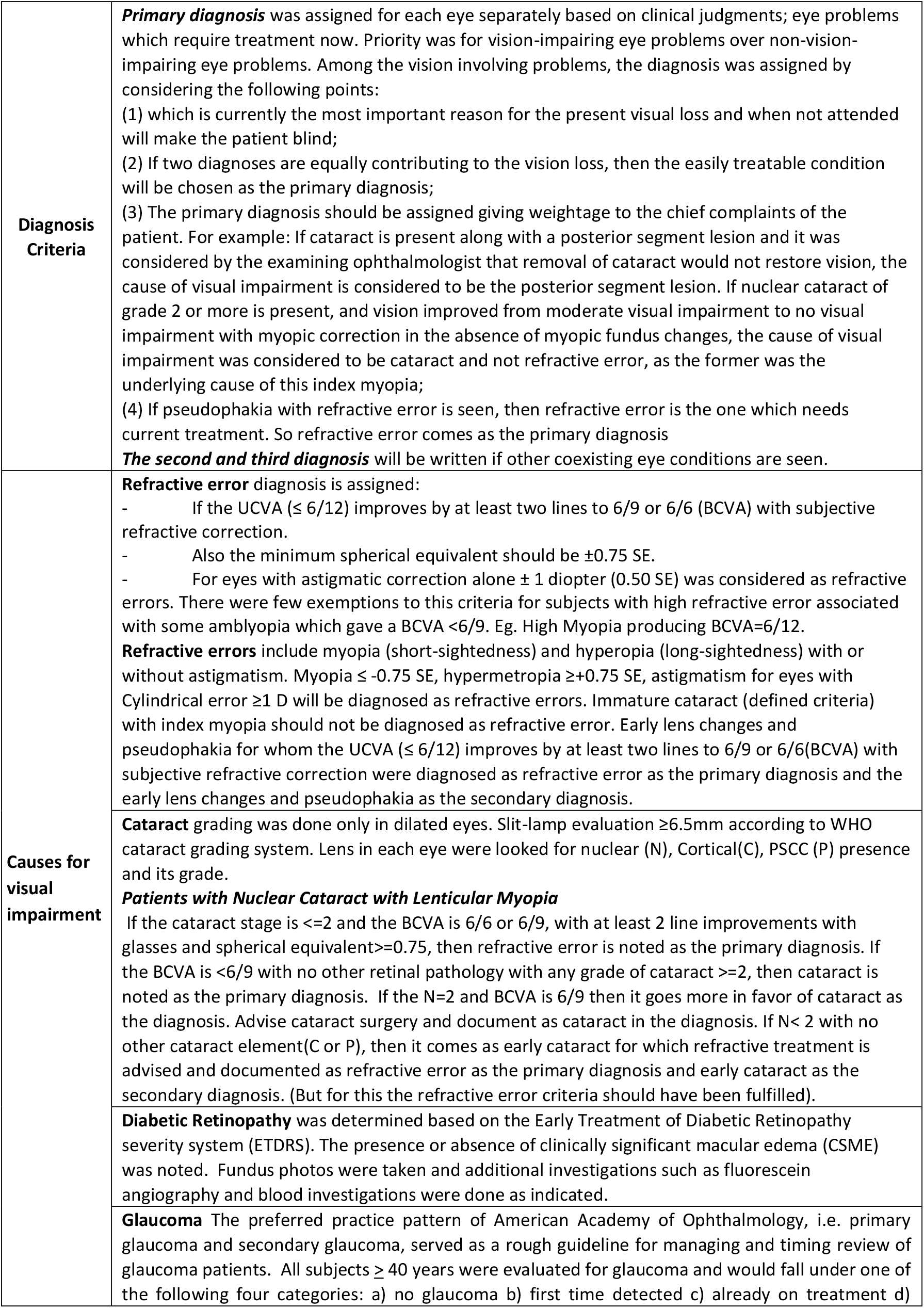
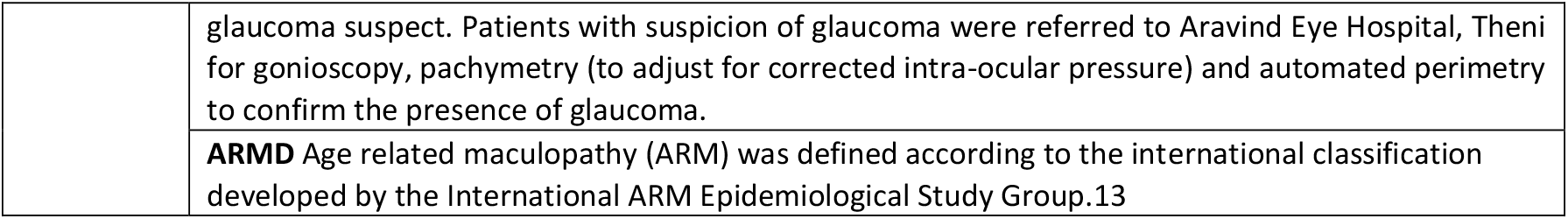
Definition of Diagnosis Criteria and Causes for Visual Impairment

|  |  |
| --- | --- |
| **Diagnosis Criteria** | **Primary diagnosis** was assigned for each eye separately based on clinical judgments; eye problems which require treatment now. Priority was for vision-impairing eye problems over non-vision-impairing eye problems. Among the vision involving problems, the diagnosis was assigned by considering the following points:<br>(1) which is currently the most important reason for the present visual loss and when not attended will make the patient blind;<br>(2) If two diagnoses are equally contributing to the vision loss, then the easily treatable condition will be chosen as the primary diagnosis;<br>(3) The primary diagnosis should be assigned giving weightage to the chief complaints of the patient. For example: If cataract is present along with a posterior segment lesion and it was considered by the examining ophthalmologist that removal of cataract would not restore vision, the cause of visual impairment is considered to be the posterior segment lesion. If nuclear cataract of grade 2 or more is present, and vision improved from moderate visual impairment to no visual impairment with myopic correction in the absence of myopic fundus changes, the cause of visual impairment was considered to be cataract and not refractive error, as the former was the underlying cause of this index myopia;<br>(4) If pseudophakia with refractive error is seen, then refractive error is the one which needs current treatment. So refractive error comes as the primary diagnosis<br>**The second and third diagnosis** will be written if other coexisting eye conditions are seen. |
| **Causes for visual impairment** | **Refractive error** diagnosis is assigned:<br>- If the UCVA ($\leq 6/12$) improves by at least two lines to 6/9 or 6/6 (BCVA) with subjective refractive correction.<br>- Also the minimum spherical equivalent should be $\pm 0.75$ SE.<br>- For eyes with astigmatic correction alone $\pm 1$ diopter (0.50 SE) was considered as refractive errors. There were few exemptions to this criteria for subjects with high refractive error associated with some amblyopia which gave a BCVA $< 6/9$. Eg. High Myopia producing BCVA=6/12.<br>**Refractive errors** include myopia (short-sightedness) and hyperopia (long-sightedness) with or without astigmatism. Myopia $\leq -0.75$ SE, hypermetropia $\geq +0.75$ SE, astigmatism for eyes with Cylindrical error $\geq 1$ D will be diagnosed as refractive errors. Immature cataract (defined criteria) with index myopia should not be diagnosed as refractive error. Early lens changes and pseudophakia for whom the UCVA ($\leq 6/12$) improves by at least two lines to 6/9 or 6/6(BCVA) with subjective refractive correction were diagnosed as refractive error as the primary diagnosis and the early lens changes and pseudophakia as the secondary diagnosis.<br>**Cataract** grading was done only in dilated eyes. Slit-lamp evaluation $\geq 6.5$mm according to WHO cataract grading system. Lens in each eye were looked for nuclear (N), Cortical(C), PSCC (P) presence and its grade.<br>**Patients with Nuclear Cataract with Lenticular Myopia**<br>If the cataract stage is $\leq 2$ and the BCVA is 6/6 or 6/9, with at least 2 line improvements with glasses and spherical equivalent $\geq 0.75$, then refractive error is noted as the primary diagnosis. If the BCVA is $< 6/9$ with no other retinal pathology with any grade of cataract $\geq 2$, then cataract is noted as the primary diagnosis. If the N=2 and BCVA is 6/9 then it goes more in favor of cataract as the diagnosis. Advise cataract surgery and document as cataract in the diagnosis. If N$< 2$ with no other cataract element(C or P), then it comes as early cataract for which refractive treatment is advised and documented as refractive error as the primary diagnosis and early cataract as the secondary diagnosis. (But for this the refractive error criteria should have been fulfilled).<br>**Diabetic Retinopathy** was determined based on the Early Treatment of Diabetic Retinopathy severity system (ETDRS). The presence or absence of clinically significant macular edema (CSME) was noted. Fundus photos were taken and additional investigations such as fluorescein angiography and blood investigations were done as indicated.<br>**Glaucoma** The preferred practice pattern of American Academy of Ophthalmology, i.e. primary glaucoma and secondary glaucoma, served as a rough guideline for managing and timing review of glaucoma patients. All subjects $\geq 40$ years were evaluated for glaucoma and would fall under one of the following four categories: a) no glaucoma b) first time detected c) already on treatment d) |
|  | glaucoma suspect. Patients with suspicion of glaucoma were referred to Aravind Eye Hospital, Theni for gonioscopy, pachymetry (to adjust for corrected intra-ocular pressure) and automated perimetry to confirm the presence of glaucoma. |
|  | **ARMD** Age related maculopathy (ARM) was defined according to the international classification developed by the International ARM Epidemiological Study Group.<sup>13</sup> |

**Figure 3:**
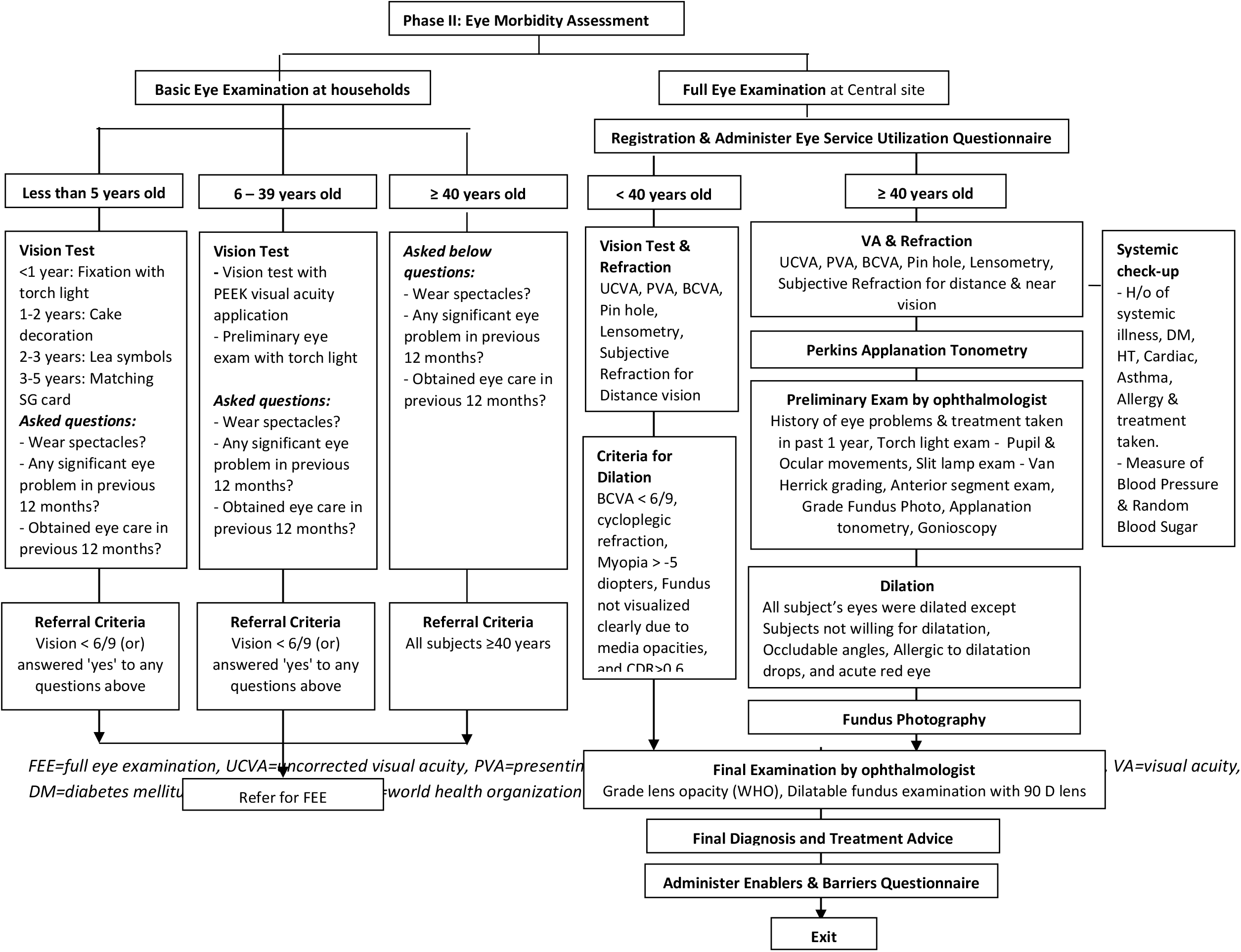
Eye morbidity assessment

### Diagnosis criteria and causes for VI

Table-2 provides details of the diagnosis criteria for selected vision impairing eye conditions. Principal cause of VI was recorded for an eye with PVA ≤6/18. Primary and secondary eye diagnoses were decided by study ophthalmologists based on the comprehensive eye examination.

### Treatment provided

Eye drops, near vision glasses, and cataract surgery were offered free of cost. A flat fee of 100 rupees was charged for dispensing bifocal/distance spectacles. All other essential treatments were provided either free of cost or at substantially subsidized rates. Those subjects requiring further investigations for glaucoma and/or retinal eye diseases were referred to AEH-Theni. Up to three phone call reminders were made to encourage them to comply with the referral. Besides phone call reminders, door-to-door awareness, travel allowance, transport facility for pickup and drop, free medicines, and spectacles were offered. All advanced investigations were done free of cost at the base hospital.

### Enablers & Barriers interview

A pre-tested, structured ‘enablers and barriers questionnaire’ was administered to the subjects by trained interviewers. For subjects aged ≤15 years, their parents/guardians were interviewed. Questions under “Enablers” were categorized as felt need, awareness, attitude, societal support, accessibility and affordability; and the “Barriers” questions were categorised as awareness, attitude, beliefs, societal support, personal/family/job obligations, economic and accessibility. For each question, the closest responses were checked from the prelisted options. In order to ensure validity of the responses, multiple responses were captured and the subjects were asked to select the single most significant factor. Each interview lasted for 10-15 minutes.

### Mop-up survey

We had an initial response rate of 54% at “FEE”. In order to improve the response rate, we conducted a mop-up survey with various interventions such as examination hours to suit the convenience of subjects, provision of medicines and spectacles free of cost and door-to-door subject mobilization by the study staff. Additional efforts included seeking support from local leaders and volunteers and arranging transport, especially, for the aged and/or sick people. Each cluster took 2 days to complete this mop-up survey. Eye examination was performed at HH for all ages due to the following instances – (a) late arrival of subjects to home after work, (b) hesitation to get eye exam at the central campsite and (c) physical illness. Distance vision was measured using PEEK VA mobile application. If uncorrected VA or PVA were worse than 6/12, then VA was measured using pinhole to assess the presence of any refractive error. Blood pressure, capillary blood sugar level and intra-ocular-pressure (Perkin’s applanation tonometer) were measured by ophthalmic assistants. This was followed by handheld slit lamp examination by ophthalmologist. Subjects needing further examination were examined using a static slit lamp set up inside a mobile van. The mop-up survey helped to enhance the overall response rate up to 72.4% in <40 years and 70.3% in ≥40 years.

### Data management and quality control

Data were collected using printed forms and were entered into a Microsoft Access 2010 based database. Double data entry was done, one in the field and the second off line to ensure data quality. To ensure accuracy and completeness, the forms were manually checked at the end of examination before the participant left the study premises. Fundus images were stored in study laptops with study IDs. Backup of the database including the images were saved to an encrypted hard disk on a daily basis. Inter-observer agreement was checked between ophthalmologists for gonioscopy, lens opacity grading, applanation tonometry, primary diagnosis and between ophthalmic technicians for VA and refraction.

### Statistical Analysis

The normality of the data was checked using Shapiro Wilk’s test. Chi square test or Fisher’s exact test was used to find the association between categorical variables. Student’s T-test and Paired T-test were used to compare the continuous variables between groups and paired comparisons. The association between the independent variables and the outcome variable was analysed using logistic regression adjusted for multiple factors. Odds ratios were computed with 95% confidence interval. *P* value <0.05 was considered as statistically significant. We used STATA-14 (Texas, USA) for statistical analysis.

## Results

We enrolled 7,386 households from which 24,327 subjects were recruited. Overall, 10,270 participants completed the BEE and 8,853 participants completed the FEE of which 1,726 had age < 40 years and 7,127 were aged 40 years or older. Among those aged <40 years, 1,132 were advised treatment and 686 completed the interview. Among those aged 40 years or above, 6,310 were advised treatment and 2,901 completed the interview. A detailed description of the recruitment and participation of the study subjects is given in Figure-4.

**Figure 4:**
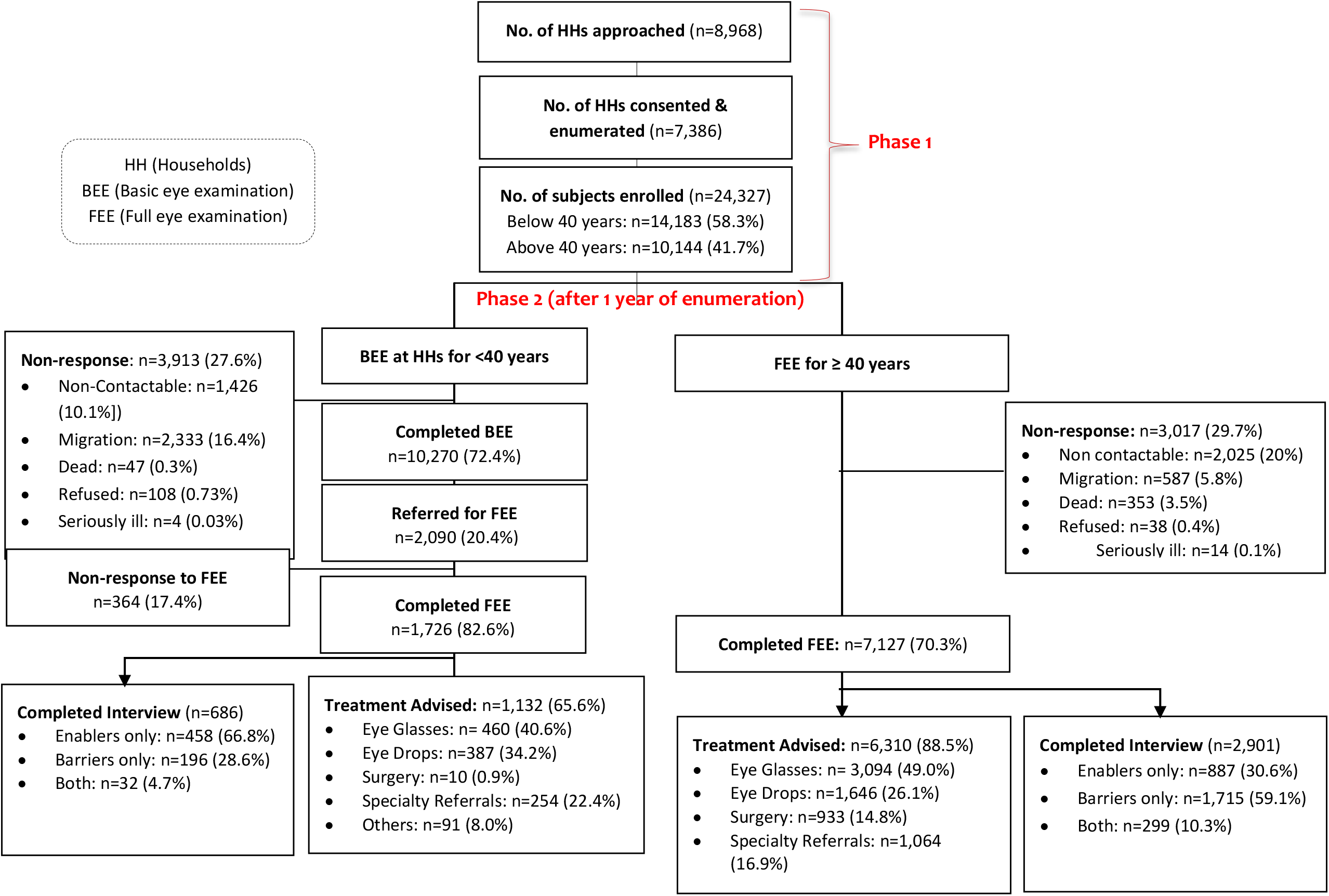
Survey Participants

## Discussion

We envisage that our study findings would enable us to develop a model to estimate the expected demand of annual eye care needs for a given population. Moreover, this might serve as a parameter for eye care providers to monitor their annual performance for a given geographic region. Further, it is possible to conduct CEWA as a cross sectional survey (without the baseline enumeration phase) in places where electronic medical records are widely used with unique patient identifiers.

The key strengths of our study methodology is the random cluster sampling strategy and a large sample size representing both urban and rural residents of Theni district. Though the measurement of VA as part of the BEE was performed by ophthalmic assistants, the use of PEEK VA app (17, 18) ensured accuracy and consistency. During enumeration, enumerators were instructed not to provide information about eye care and services available so that the natural health seeking behaviour of people was not influenced. We believe that the use of differential protocols [1. basic eye examination at the households for people aged <40 years and 2. comprehensive eye examination at a central site within each cluster for those aged 40 years and above] made the eye examination process efficient and cost effective compared to performing comprehensive eye examinations for all age groups.

One of the study limitations was the inability to track and validate the utilization by subjects who accessed services from providers other than AEH. However, we do not foresee this to have any significant influence on our findings because AEH, through its 9 centres provides more than 80% of all eye care in Theni district. Another limitation was the comparatively low response rate in people ≥40 years. This was mainly due to non-availability of the subjects during follow-up survey because of their work timings, migration or death. During enumeration, our team could not physically cross verify the number of people in each household provided by the contact person. This, to some extent could have resulted in the non-availability of some of the members when we approached for the follow-up survey.

In conclusion, the study findings can be used for appropriate long-term public health intervention planning, resource allocation, efficient delivery of care, and designing of eye care services in similar resource-limited settings.

## Data Availability

All data produced in the present study are available upon reasonable request to the authors

## Author contributions

TR conceptualised the study and oversaw the execution of the study; TR, AV, SJ and SKR developed the study design. AEF provided guidance in the study design and implementation and provided feedback on the manuscript. AV, VKR, DD and SKR coordinated the field work and data collection. LP performed the statistical analysis and AV, SJ and VKR drafted the manuscript with guidance from TR. All authors reviewed and finalised the manuscript.

